# Translation and Cross-cultural Validation of Leprosy Case Detection Delay Questionnaire Among Persons Affected by Leprosy in Southeast Nigeria

**DOI:** 10.64898/2026.06.06.26355058

**Authors:** Chinwe Eze, Ngozi Murphy-Okpala, Ngozi Ekeke, Charles Nwafor, Daniel Egbule, Martin Njoku, Okechukwu Ezeakile, Anthony Meka, Francis S. Iyama, Emmanuel Ogbuefi, Onyeka Ugwu, Miriam Solomon, Clement Adesigbin, Joseph Chukwu

**Author notes:** **Corresponding author:** Chinwe Eze., Email address (CE).

## Abstract

**Introduction:** Reducing delays in leprosy case detection is essential for achieving global leprosy targets. Accurate measurement of these delays and their determinants relies largely on patient-reported data, as routine health records are often inadequate. The leprosy case detection delay (CDD) questionnaire, developed under the Post Exposure Prophylaxis for Leprosy (PEP4LEP) project, has been validated in Ethiopia, Mozambique, Tanzania, and Indonesia. However, it has not been adapted or validated for Nigeria or any major Nigerian indigenous language. This study aimed to culturally adapt and validate the CDD questionnaire for Igbo-speaking populations in Nigeria.

**Methodology/Principal Findings:** The CDD questionnaire underwent a standardized cross-cultural adaptation process. Content validity was assessed using item- and scale-level content validity indices, while construct validity was evaluated through hypothesis testing. Reproducibility was assessed using test–retest and inter-rater reliability; agreement using the Bland–Altman method and the Wilcoxon Signed-Rank test; reliability using Spearman’s rank correlation coefficient and the Intraclass Correlation Coefficient (ICC); and internal consistency using Cronbach’s alpha. Data were collected through face-to-face interviews with persons affected by leprosy at two time points separated by at least two weeks.

Participants (n=100) had a mean age of 45.1 years (SD=18.7). Mean CDD was 77.2 months at baseline and 77.9 months at retest. The instrument demonstrated excellent content validity (I-CVI/S-CVI: 0.90–1.00), good internal consistency (Cronbach’s α=0.77), and excellent test–retest reliability (ICC=0.996, 95% CI: 0.994–0.997). Test and retest measurements were highly correlated (ρ=0.985, p<0.001), with no evidence of systematic change over time (p=0.864). Seventy-two percent of participants reported identical CDD values across assessments. All items from the original English version were retained without modification.

**Conclusion/Significance:** The Igbo version of the CDD questionnaire demonstrated good validity and reliability and is suitable for assessing leprosy case detection delay among Igbo-speaking populations in Nigeria

## Introduction

Leprosy is a chronic infectious disease caused by *Mycobacterium leprae* and remains one of the leading causes of permanent deformity and disability attributable to a single infectious agent (1). It is one of the 21 neglected tropical diseases (NTDs) recognized by the World Health Organization (WHO) and is targeted for elimination (2). The introduction and scale-up of multidrug therapy (MDT) marked a significant breakthrough in leprosy control, resulting in a sustained global decline in new cases from over 700,000 in 2000 to 172,717 in 2024 (3). A similar trend has been observed in Nigeria, where reported new cases declined from 3,623 in 2011 to 1,770 in 2024, although pockets of high endemicity persist in some states (4).

Leprosy primarily affects the skin and peripheral nerves. When diagnosis and treatment are delayed, progressive nerve damage can result in permanent deformities and disability. Visible deformities, classified as leprosy Grade 2 Disability (G2D), are associated with stigma, discrimination, reduced social participation, mental health challenges, and economic loss among persons affected by leprosy (5–7). Consequently, G2D has gained prominence as a more robust indicator of leprosy control performance than prevalence alone. In line with this, the WHO targets a 90% reduction, from the 2020 baseline, in the rate per million population of new leprosy cases presenting with G2D as a key elimination milestone (8).

The hallmark of leprosy control is early detection and treatment. New cases with G2D indicate delayed case detection. The global trend for leprosy G2D rates per million population shows a concerning paradox. While overall new leprosy cases are declining, the number and rate per million of new cases presenting with G2D have generally increased from the 2020 figure, indicating persistent challenges with late diagnosis and treatment (3). Similarly, Nigeria has reported a G2D percentage rate among new cases of 10-15%, persistently higher than the global average of 5-7% since 2020 (3), despite the declining incidence of leprosy.

Addressing delays in leprosy case detection is critical to achieving the global leprosy targets, and this starts with quantifying the extent of the delay and identifying the associated factors. Detection delay is typically conceptualized as two components: patient delay (the time from symptom onset to first care-seeking) and health system delay (the time from first care-seeking to definitive diagnosis). In many endemic settings, routine health records are insufficient for this purpose, making patient-reported data essential.

The leprosy case detection delay questionnaire is a structured tool, developed under the Post Exposure Prophylaxis for Leprosy (PEP4LEP) project in Ethiopia, Mozambique, and Tanzania, to determine the duration of detection delays in leprosy. The tool captures the interval between the onset of initial signs and symptoms and confirmed diagnosis, the nature and location of early symptoms, care-seeking actions, number of healthcare visits, and uses locally adapted calendars and visual aids to improve recall accuracy (9). The developers recommended its use in other endemic countries and cultural settings(9).

However, in line with best practice in measurement science, health assessment instruments must undergo cross-cultural adaptation and validation before use in new settings to ensure reliability, validity, and cultural relevance (10). Without such validation, results may be misleading, inaccurate, and incomparable across different populations. The CDD questionnaire has been successfully adapted and validated in the original PEP4LEP countries and subsequently in Indonesia, demonstrating good reliability and validity, and ease of administration without undue burden on respondents (9,11,12). To date, no similar validation has been conducted in the Nigerian context or in any of the country’s major indigenous languages.

Nigeria is linguistically and culturally diverse, comprising six geopolitical zones and three major indigenous languages—Igbo, Hausa, and Yoruba. Igbo is predominantly spoken in the Southeast and parts of the South-South regions. In this diverse setting, direct translation of health questionnaires is insufficient; concepts of illness, local nomenclature for skin conditions, and traditional health-seeking behaviors must be culturally adapted to ensure that questions are accurately interpreted and that the data collected are reliable and relevant to the Nigerian population. This study, therefore, aimed to translate the CDD questionnaire into Igbo, cross-culturally adapt it, and assess its validity and reliability for use in the Nigerian context.

## Methods

### Study setting

This study was conducted in Ebonyi and Enugu States (Fig 1), two of the five states in Nigeria’s southeastern region, where Igbo is the predominant language. Ebonyi and Enugu States comprise 13 and 17 local government areas (LGAs), respectively, with projected 2024 populations of 3,591,241 and 5,563,273.

**Fig 1:**
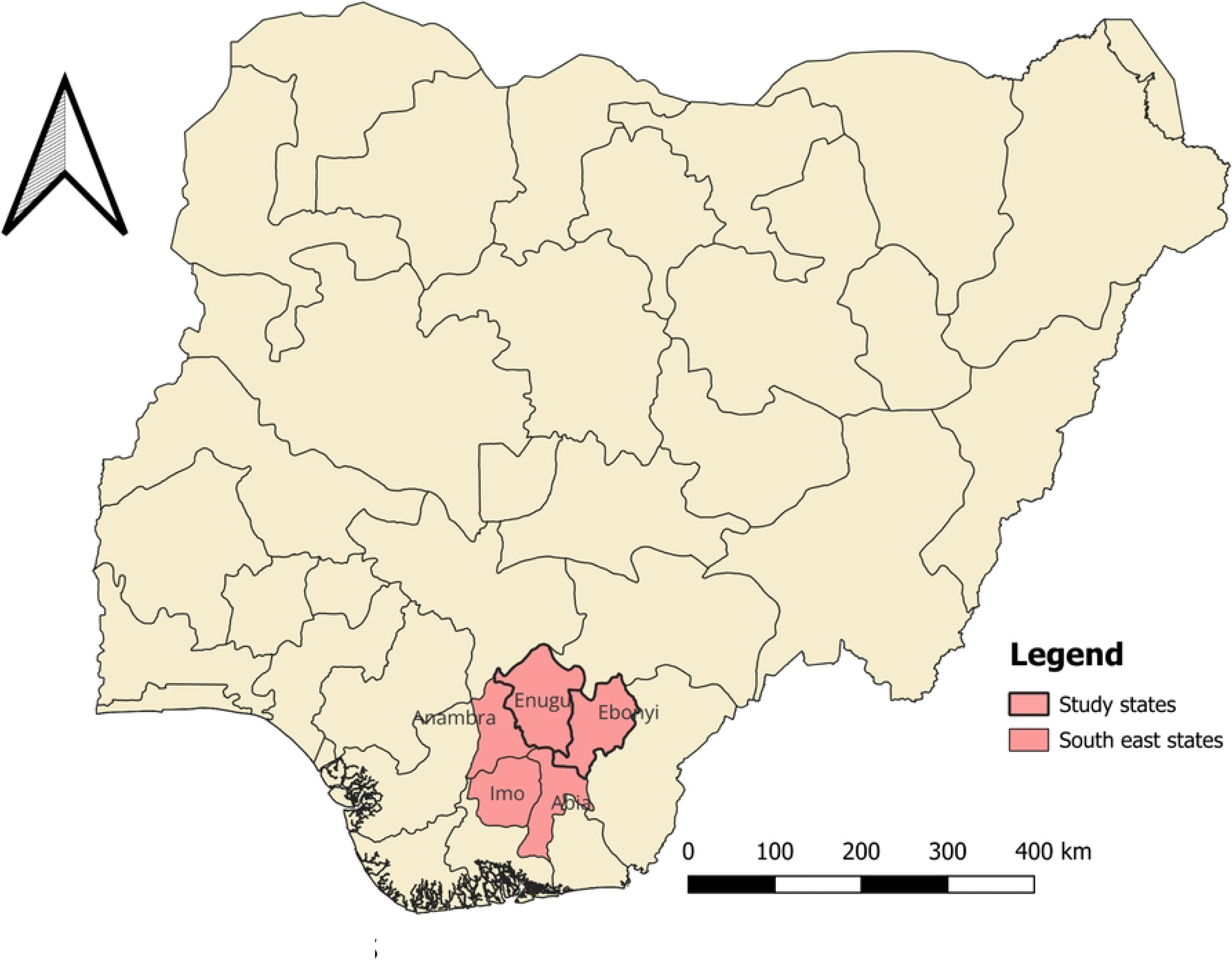
Map of Nigeria showing Southeast states.

Agriculture, particularly subsistence farming, is the main livelihood in both states, although trading and service-based occupations are common in the more urban areas of Enugu State.

Ebonyi and Enugu States rank first and second, respectively, among the southeastern states in terms of five-year cumulative new leprosy case notification and Grade 2 Disability (G2D). In 2024, of the 174 new leprosy cases and 61 cases with G2D reported in the region, the two states accounted for 78% (136) of new cases and 74% (45) of G2D cases.

Leprosy control in Nigeria is primarily implemented at the LGA level, with Local Government Tuberculosis, Buruli Ulcer, and Leprosy Supervisors (LGTBLS) responsible for case detection, diagnosis, treatment initiation, and treatment monitoring. Each of the two states has one designated leprosy referral facility and a few LGAs that account for more than 70% of their annual new cases-approximately three in Ebonyi State and two in Enugu State.

### Study design

A cross-sectional design was employed.

### Study population

The study population comprised persons affected by leprosy who were either currently on treatment or had been released from treatment and who provided informed consent to participate. To minimize recall bias, individuals who had been recently diagnosed were prioritized. Persons who were severely ill or had mental conditions that could impair their ability to provide reliable information were excluded from the study.

### Sampling and Sample Size

There are no universally agreed-upon rules for sample size determination in questionnaire validation studies (13). However, a commonly applied rule of thumb recommends recruiting between three and ten respondents per questionnaire item (14,15). The CDD questionnaire comprises 10 items. Using an average of 5 respondents per item and allowing for a 20% attrition rate to accommodate potential loss to follow-up in the test–retest reliability assessment, a minimum sample size of 63 participants was estimated.

To ensure sufficient statistical power and enhance the robustness of the reliability estimates, a total of 100 respondents were finally recruited. Sampling was conducted in LGAs with the highest leprosy case notification in each state. Eligible participants were identified through leprosy treatment registers and invited to participate in the study; those who consented and were available at the time of data collection were enrolled.

### Data collection and analysis

Participant recruitment and data collection spanned from March to June 2025. Data were collected through face-to-face interviews conducted by trained LGTBLSs. The use of LGTBLSs was necessary because of their familiarity with leprosy case detection, diagnosis, and treatment, which facilitated accurate administration of the questionnaire and appropriate probing where necessary. Before data collection, interviewers received standardized training on the study protocol, ethical considerations, and administration of the leprosy case detection delay (CDD) questionnaire. The accompanying question-by-question guide to the CDD questionnaire was used during the training, and a copy was given to each interviewer afterward for reference.

The finalized questionnaire was deployed via the CommCare application (Dimagi Inc.) on Android phones. CommCare is an electronic data capture platform that supports case management and validation checks. Digitizing data collection for this study enabled real-time monitoring of data completeness, consistency, and prompt resolution of identified issues, thereby enhancing data quality.

Data were exported from CommCare into IBM SPSS Statistics version 25 for analysis. Descriptive statistics were used to summarize participants’ sociodemographic and clinical characteristics. Continuous variables were assessed for normality using visual inspection and summary statistics. Given the skewed distribution of case detection delay, appropriate non-parametric and transformed analyses were applied as described in the reliability and agreement assessments. Test–retest reliability and inter-rater reliability were evaluated using Spearman’s rank correlation coefficient and the Intraclass Correlation Coefficient (ICC), respectively. Agreement between baseline and retest measurements was further assessed using Bland–Altman analysis. All statistical tests were two-sided, and p-values <0.05 were considered statistically significant.

### Measurements

#### Content validity and cross-cultural adaptation

The leprosy CDD questionnaire underwent a standardized cross-cultural adaptation process (Fig 2)(16). Two independent bilingual linguistic lecturers from Ebonyi State University translated the original English questionnaire into Igbo. The two forward translations were reviewed by the Principal Investigator (PI) in a consensus meeting with the translators, during which discrepancies were discussed and resolved, resulting in a single reconciled Igbo version.

**Fig 2:**
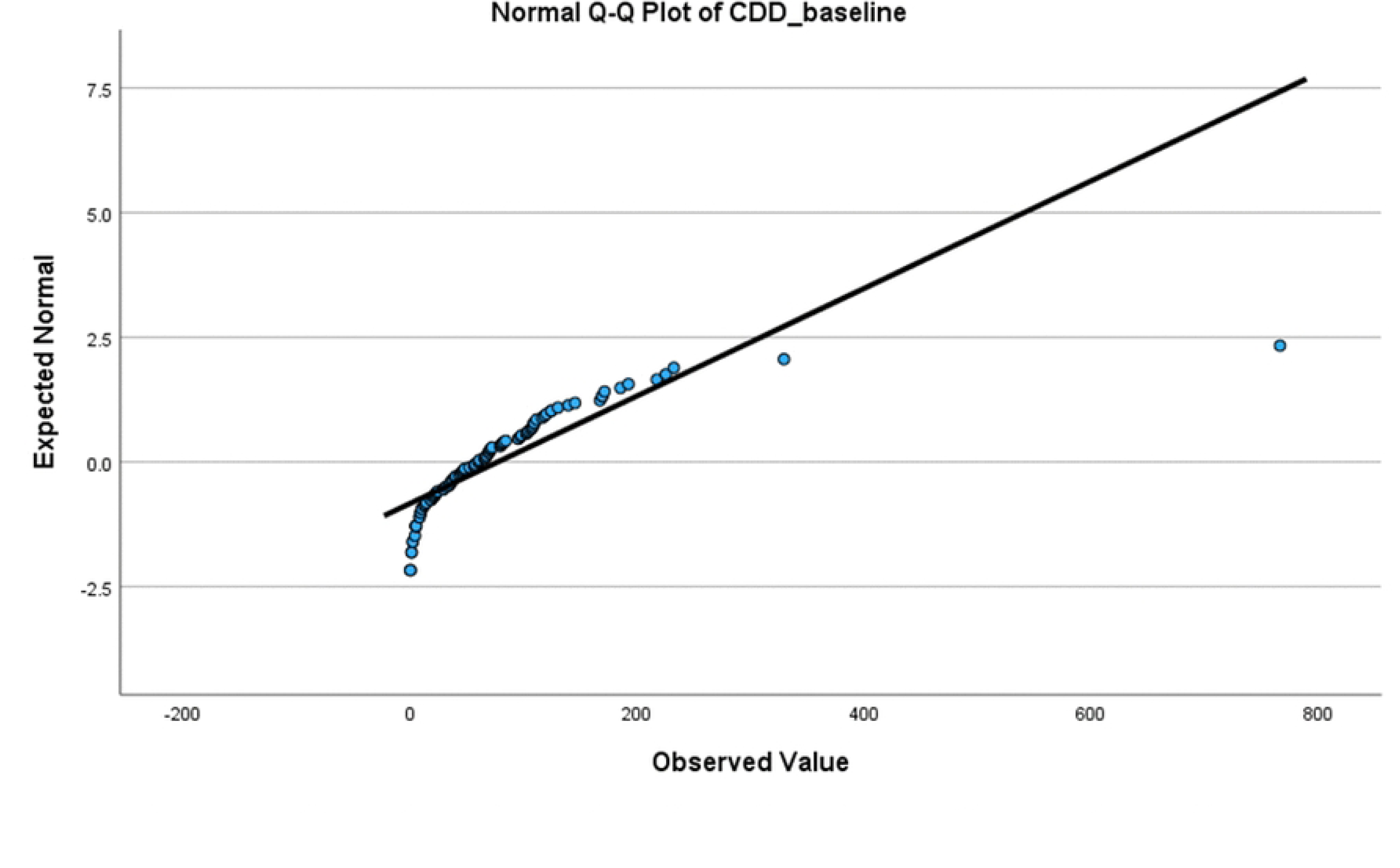
Flow chart translation and cross-cultural adaptation process.

The reconciled Igbo version was subsequently back-translated into English by a separate pair of independent bilingual linguistic lecturers from the same institution who had not been involved in the forward translation. The back-translated version was compared with the original English questionnaire, and discrepancies were reviewed and resolved by consensus, resulting in a finalized Igbo version. Throughout the translation process, translators were blinded to each other’s work until the review stage to minimize bias.

An expert committee comprising four public health physicians with extensive experience in leprosy management and program implementation (mean experience of 20 years) and one person affected by leprosy reviewed the translated instrument. The committee assessed conceptual, semantic, and qualitative item equivalence by comparing the original English version, the Igbo translation, and the back-translated English version. Minor modifications were made to improve clarity and cultural relevance, and the Igbo version was deemed equivalent to the original questionnaire.

Item equivalence was further assessed quantitatively using a five-point Likert-scale questionnaire administered to seven public health physicians involved in leprosy control programs. Experts rated each CDD questionnaire item for relevance and clarity and assessed the instrument’s overall comprehensiveness in measuring the delay in leprosy case detection. Ratings were provided on a five-point Likert scale ranging from 1 (not relevant / not clear / not comprehensive) to 5 (highly relevant / very clear / very comprehensive). Ratings of 4 or 5 were considered adequate. Item-level Content Validity Index (I-CVI) was calculated as the proportion of experts rating an item 4 or 5. Values ≥ 0.83 were considered valid for relevance or clarity(17). Scale-level CVI (S-CVI/Ave) was computed as the average of I-CVI values across all items. S-CVI ≥ 0.8 is considered valid and <0.78, invalid (17).

Operational equivalence was assessed through cognitive interviews conducted by a trained LGTBLS using the instrument’s pre-final version. Ten individuals affected by leprosy were interviewed to evaluate clarity, comprehension, and acceptability. Feedback indicated no need for further modifications and confirmed that face-to-face interviews were the most appropriate mode of administration, particularly given the limited literacy levels among respondents.

#### Construct Validity

Construct validity was assessed using a hypothesis-testing approach, as no validated reference instrument for measuring CDD exists in the Nigerian context. This was adopted from a similar study in Indonesia (11). This approach is consistent with recommended methods for evaluating construct validity when criterion validity cannot be assessed, particularly in the absence of a gold-standard reference instrument (15). The following hypotheses, adapted from the Indonesian study, were tested:

1. The calculated case detection delay should yield a positive value, reflecting the logical expectation that the time from onset of symptoms to diagnosis cannot be negative.
2. Responses to the component questions used to define CDD, namely, the duration since first noticing leprosy signs or symptoms, the duration since first visiting a health facility, the duration since receiving a leprosy diagnosis, and the calculated CDD duration, should be positively correlated, as these variables represent sequential events along the care-seeking pathway.

Construct validity was considered adequate if the data supported at least 75% of the predefined hypotheses (15).

Spearman’s rank correlation coefficient was used to test the hypothesized associations because the delay variables were non-normally distributed. Correlation coefficients were interpreted in terms of strength and direction to determine whether observed relationships were consistent with theoretical expectations of the CDD construct. The proportion of supported hypotheses was calculated to determine overall construct validity.

#### Reliability

The instrument’s reproducibility was assessed using test–retest and inter-rater reliability. Reliability was examined using Spearman’s rank correlation coefficient and the ICC for agreement.

The CDD questionnaire was administered to 100 participants on two occasions separated by a minimum interval of two weeks. Spearman’s rank correlation coefficient was used to assess the test–retest association because the delay data were skewed. Spearman’s rho values were interpreted as follows: approximately 0.30 indicating weak correlation, 0.50 moderate, 0.70 strong, and ≥0.90 very strong correlation (15).

Different raters were trained and independently administered the questionnaire at each time point. Consequently, the ICC estimates reflect both temporal stability (test–retest reliability) and agreement between raters (inter-rater reliability). ICC was estimated using a two-way mixed-effects model with absolute agreement for single measurements on log-transformed data, which is appropriate when the same participants are assessed by different raters and the interest is in exact agreement rather than consistency alone. ICC values were interpreted according to established criteria: values <0.70 indicate poor reliability, 0.70–0.79 moderate reliability, 0.80–0.89 good reliability, and ≥0.90 excellent reliability (18).

#### Internal Consistency

Internal consistency refers to how well the set of questions in an instrument is correlated, thus measuring the same construct (15). We measured it using Cronbach’s alpha. Internal consistency is considered good if the value ranges from 0.7 to 0.95 (15). Four items of the CDD questionnaire that directly measure the duration of case detection delay were assessed for internal consistency (Box 1). Confirmatory items were excluded from the reliability coefficient calculations.

###### Box 1: Questionnaire Items related to case detection delay

1. To specify, how many months ago did you notice the first signs of your disease?
2. When was your first visit to a health facility? (How many months ago)
3. When did you receive your diagnosis of leprosy? (How many months ago)
4. Calculation of delay: subtract the time of initial sign onset from the time when the person was diagnosed as a leprosy patient (in months)

#### Test of agreement

Agreement between baseline and retest CDD questionnaire measurements was assessed using the Wilcoxon Signed-Rank test and the Bland–Altman method. A non-statistically significant Wilcoxon Signed-Rank test value indicates the absence of systematic bias between the baseline and retest measurements.

Similarly, the Bland-Altman test evaluates absolute agreement by examining the distribution of differences between paired measurements rather than their correlation (19). For each participant, the difference between baseline and retest scores was plotted against the mean of the two measurements. The mean difference (bias) and the 95% limits of agreement (mean difference ±1.96 standard deviations) were calculated to quantify systematic bias and the range within which most differences between repeated measurements are expected to lie. Visual inspection of the Bland–Altman plot was used to assess the presence of systematic bias, trends related to the magnitude of delay (proportional bias), and outliers.

Given the skewed distribution of case detection delay, Bland–Altman analysis was performed on log-transformed values. Mean differences and limits of agreement were back-transformed by exponentiation to express agreement on the original (ratio) and relative (%) scales. Agreement was considered acceptable if the mean difference was close to zero and the majority of observations lay within the 95% limits of agreement, with no evidence of proportional bias across the range of measurements.

### Ethical approval

Ethical approval was obtained from the Health Research and Ethics Committee of the University of Nigeria Teaching Hospital, Enugu. Permission was obtained from the respective states’ Tuberculosis, Leprosy and Buruli Ulcer Control Programs. Written informed consent was obtained from all respondents. Where a child was involved, assent was obtained from the child, and the parents provided written permission. All respondents were provided with study information, including details on leprosy, the study purpose, the right to withdraw, and the confidentiality of their disease status.

## Results

### Characteristics of respondents

Table 1 shows the characteristics of the respondents. A total of 100 respondents who completed both the baseline and retest assessments were included in the final analysis. Participants’ ages ranged from 6 to 80 years, with a mean age of 45.1 years (SD = 18.7; 95% CI: 41.4–48.8). Females constituted 56% of the study population, and the majority of respondents were aged ≥15 years (95%). Most respondents had multibacillary leprosy (95%).

**Table 1:**
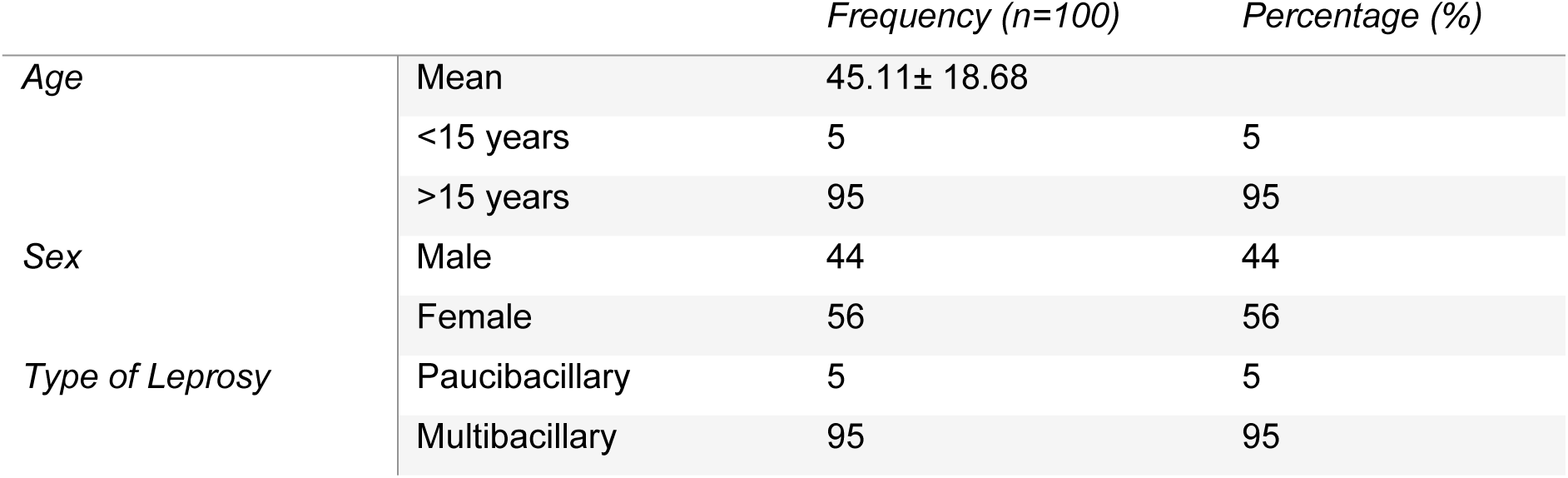
Characteristics of respondents.

|  |  | Frequency (n=100) | Percentage (%) |
| --- | --- | --- | --- |
| Age | Mean | 45.11± 18.68 | |
|  | <15 years | 5 | 5 |
|  | >15 years | 95 | 95 |
| Sex | Male | 44 | 44 |
|  | Female | 56 | 56 |
| Type of Leprosy | Paucibacillary | 5 | 5 |
|  | Multibacillary | 95 | 95 |

### Description of outcome variables

At baseline, the mean CDD was 77.2 months (SD = 92.7), with a median of 60 months (IQR: 86). At retest, the mean and median CDD were similar, at 77.9 months (SD = 91.7) and 61.5 months (IQR: 85), respectively (Table 2). The distributions of CDD at baseline and retest were positively skewed (Table 2, Fig 3, Fig 4).

**Fig 3:**
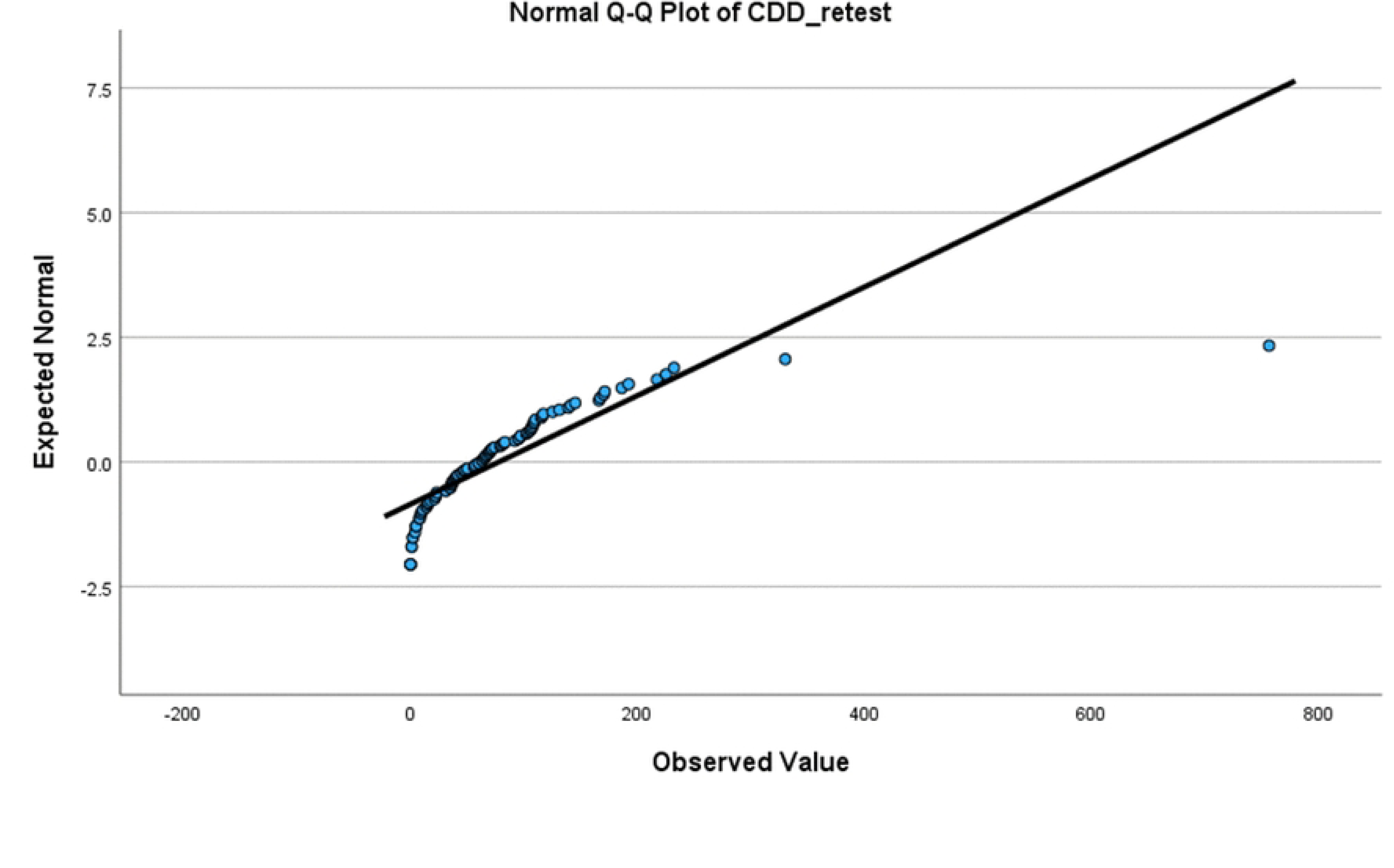
Normal Q-Q plot showing the distribution of CDD baseline scores.

**Fig 4:**
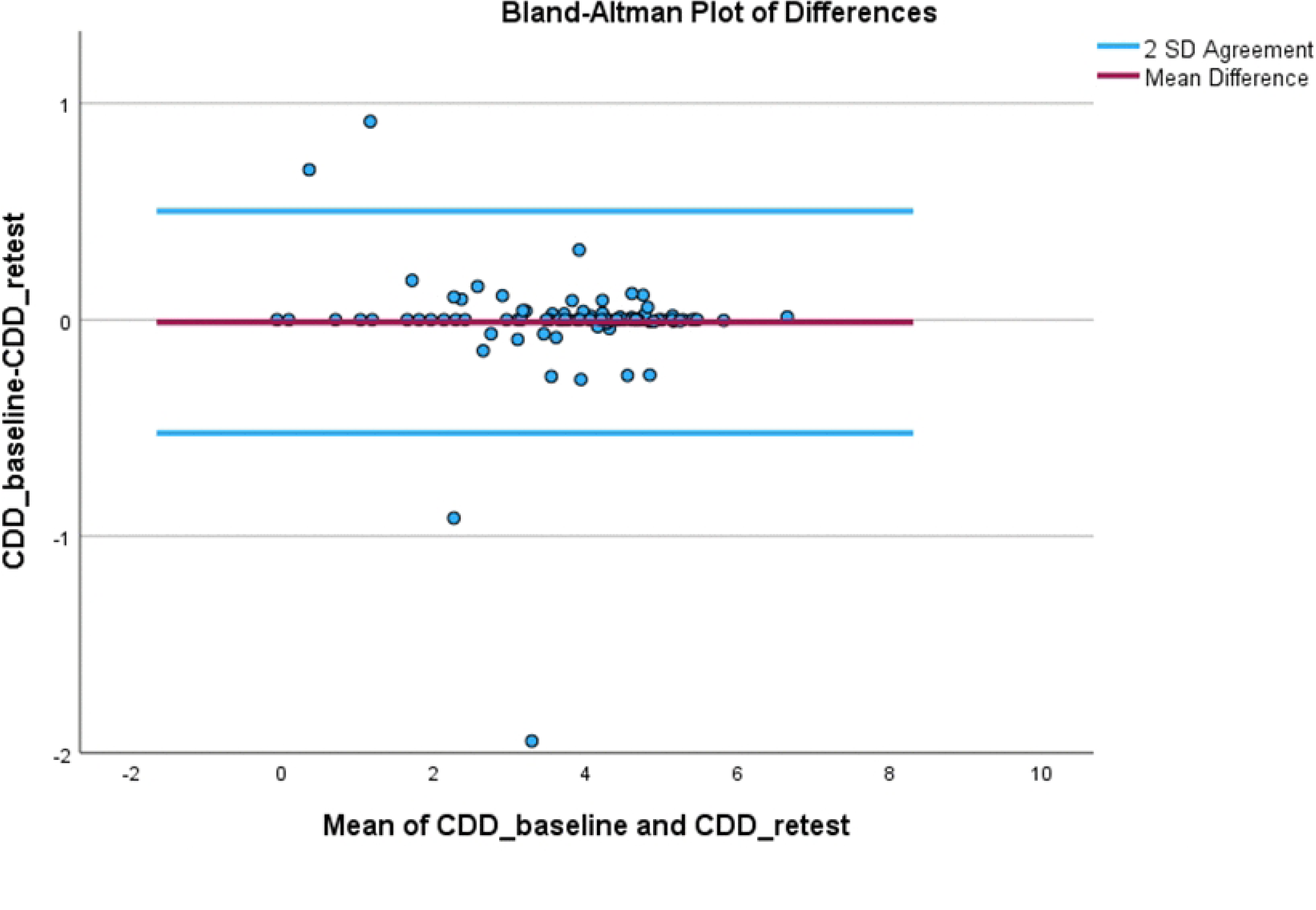
Normal Q-Q plot showing the distribution of CDD retest scores.

**Table 2:**
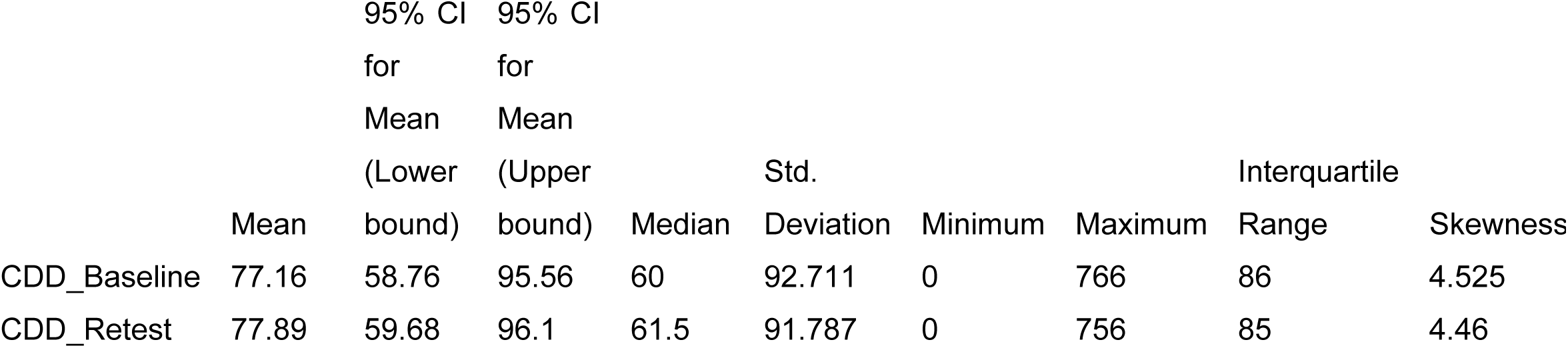
Descriptive statistics for outcome variables.

### Content validity

Feedback from the expert committee and participants involved in the pre-testing resulted in minor revisions to the Igbo version of the questionnaire, primarily to improve wording clarity, without altering item meaning. All identified discrepancies were resolved through consensus. Following these revisions, all items in the questionnaire were judged to be relevant, clear, and culturally appropriate for Igbo-speaking persons affected by leprosy in southeast Nigeria.

In the quantitative assessment (I-CVI demonstrated very good clarity (range: 0.90–1.00) for all items, except item 9, and high relevance across all items (range: 0.90–1.00). The lower clarity score for item 9 was attributed to feedback from a single expert and was subsequently clarified during the review process; however, no modification to the item wording was deemed necessary. The overall comprehensiveness index was 1.00, further supporting the tool’s strong content validity.

The S-CVI/Av), calculated as the average of item-level CVIs, was 0.98 for relevance and 0.96 for clarity, exceeding the recommended threshold of 0.90 and indicating excellent content validity of the instrument.

Consequently, all items from the original English version were retained in the Igbo version without changes to content, format, or scoring.

Table 3), I-CVI demonstrated very good clarity (range: 0.90–1.00) for all items, except item 9, and high relevance across all items (range: 0.90–1.00). The lower clarity score for item 9 was attributed to feedback from a single expert and was subsequently clarified during the review process; however, no modification to the item wording was deemed necessary. The overall comprehensiveness index was 1.00, further supporting the tool’s strong content validity.

**Table 3:** Item Content validity Index.

| Item No | Description | Relevance | Clarity |
| --- | --- | --- | --- |
| 1 | First sign noticed | 1.0 | 1.0 |
| 2 | Year first sign was noticed | 1.0 | 1.0 |
| 3 | When first sign was noticed (months) | 1.0 | 0.9 |
| 4a. | How the disease developed | 1.0 | 1.0 |
|  | Which part of the body was the first sign |  |  |
| 4b. | noticed | 0.9 | 1.0 |
| 5 | Steps taken on observing the first sign | 1.0 | 1.0 |
| 6 | First visit to health facility (in months) | 1.0 | 1.0 |
| 7 | Number of facility visits before diagnosis | 1.0 | 1.0 |
| 8 | When diagnosis was made (in months) | 1.0 | 1.0 |
| 9 | Confirmation of question 3 | 0.9 | 0.7 |
| 10 | Calculation of delay | 1.0 | 1.0 |

The S-CVI/Av), calculated as the average of item-level CVIs, was 0.98 for relevance and 0.96 for clarity, exceeding the recommended threshold of 0.90 and indicating excellent content validity of the instrument.

Consequently, all items from the original English version were retained in the Igbo version without changes to content, format, or scoring.

### Construct Validity

The median case detection delay (CDD) was 60 months (IQR: 21-107; minimum: 0; maximum: 766). The Wilcoxon Signed-Rank test showed that the distribution of CDD was significantly greater than zero (p < 0.001) (Table 4). This confirms the hypothesis that CDD values are systematically positive, indicating that individuals in this sample generally experience a measurable delay between symptom onset and diagnosis.

**Table 4:**
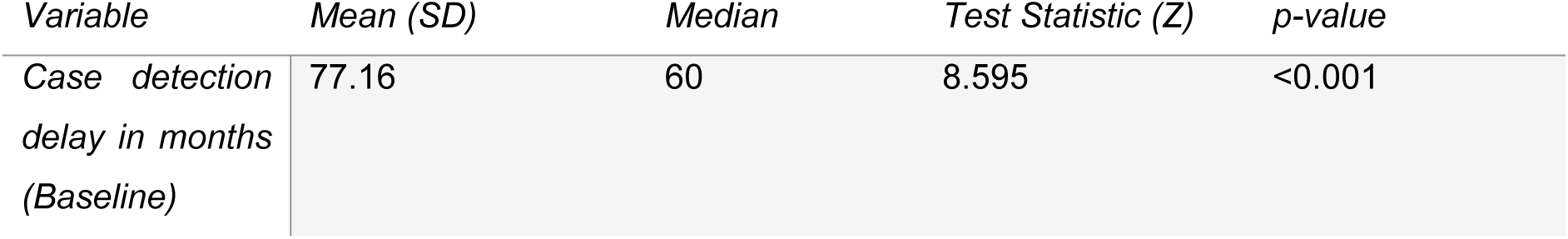
Hypothesis 1 test summary: Wilcoxon Signed-Rank Test.

The second hypothesis (that responses to all questions defining the CDD should be positively correlated) was tested using the Spearman rank correlation test. It showed a positive intercorrelation among all items of the questions defining CDD (Table 5), confirming the hypothesis.

**Table 5:**
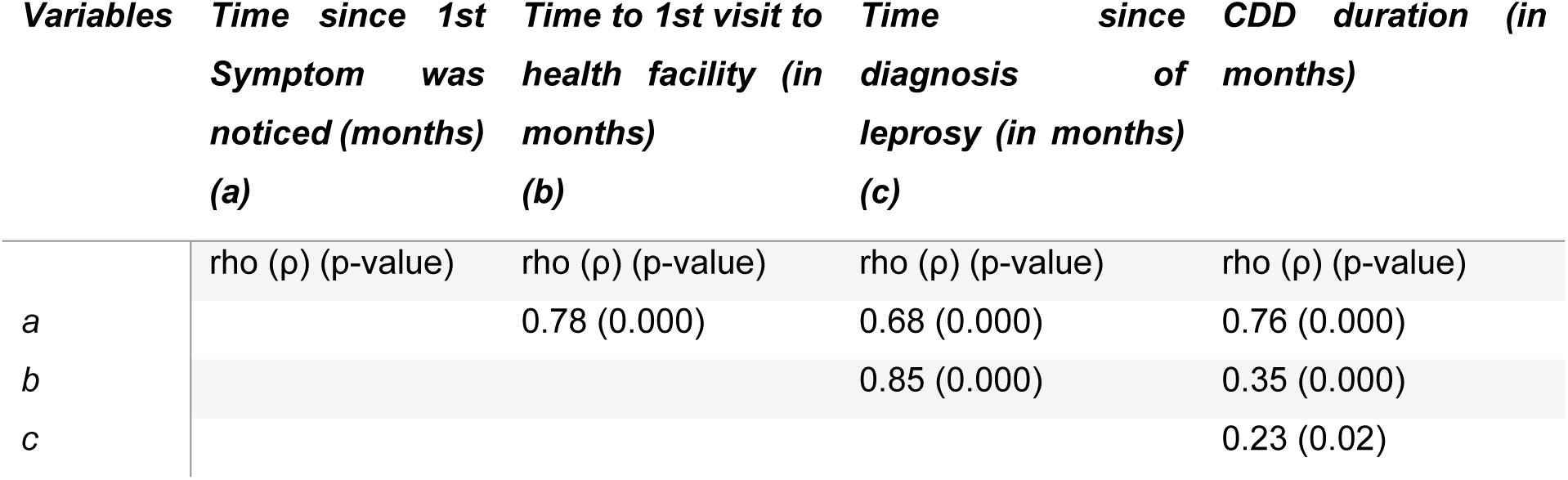
Hypothesis 2 testing summary: Rank Spearman’s correlation matrix of the main questions to determine CDD of leprosy (baseline)

### Internal Consistency

The CDD questionnaire demonstrated good internal consistency at baseline (Cronbach’s α = 0.77) and at retest (Cronbach’s α = 0.77), indicating that the items reliably measured leprosy case detection delay (Table 6).

**Table 6:** Internal consistency reliability of the CDD questionnaire.

| **Administration** | **Cronbach's $\alpha$** | **Standardized $\alpha$** | **Number of items** |
| --- | --- | --- | --- |
| *Baseline* | 0.768 | 0.775 | 4 |
| *Retest* | 0.766 | 0.774 | 4 |

### Reliability testing result (Reproducibility)

The Spearman’s rank order test showed a very strong, positive correlation between the test and retest values, which was statistically significant (ρ = .985, p < .001). The reliability of the case detection delay measurements was assessed using a two-way random effects model (absolute agreement). The ICC for single measures was .996 (95% CI: .994–.997), and for average measures was .998 (95% CI: .997–.999), F (99, 99) = 527.74, p < .001 (Table 7).

**Table 7:** Reliability and Consistency of Delay Measurements.

| **Measurement Pair** | **Spearman's $\rho$** | **Cronbach's $\alpha$** | **ICC (Single)** | **ICC (Average)** | **p-value** |
| --- | --- | --- | --- | --- | --- |
| CDD_baseline vs. CDD_retest | .985 | .998 | .996 | .998 | < .001 |

### Test of agreement

A Wilcoxon Signed-Rank test was performed to evaluate the presence of systematic change between baseline and re-test; no evidence of systematic change was found (Z = -0.172, p = .864) (Table 8). A high level of stability was observed, with 72% of the sample (n=72) recording identical values across both measurements.

**Table 8:** Systematic bias testing using the Wilcoxon-Signed Rank test.

| **Comparison** | **Negative Ranks (n)** | **Positive Ranks (n)** | **Ties (n)** | **Z-statistic** | **p-value** |
| --- | --- | --- | --- | --- | --- |
| CDD_baseline – CDD_retest | 18 | 10 | 72 | -.172 | 0.864 |

Bland–Altman analysis of the natural log-transformed baseline (CDD1) and retest (CDD2) scores showed a mean difference (bias) of −0.011 log units (95% CI: −0.062 to 0.040). This is equivalent to a 1.1% difference on the natural scale, indicating no statistically significant systematic bias between the two measurements. The 95% limits of agreement ranged from −0.524 to 0.501 log units (95% CI: −0.612 to −0.436 and 0.413 to 0.590), suggesting that approximately 95% of individual differences between CDD_baseline and CDD_retest lay within these bounds (Table 9).

**Table 9:**
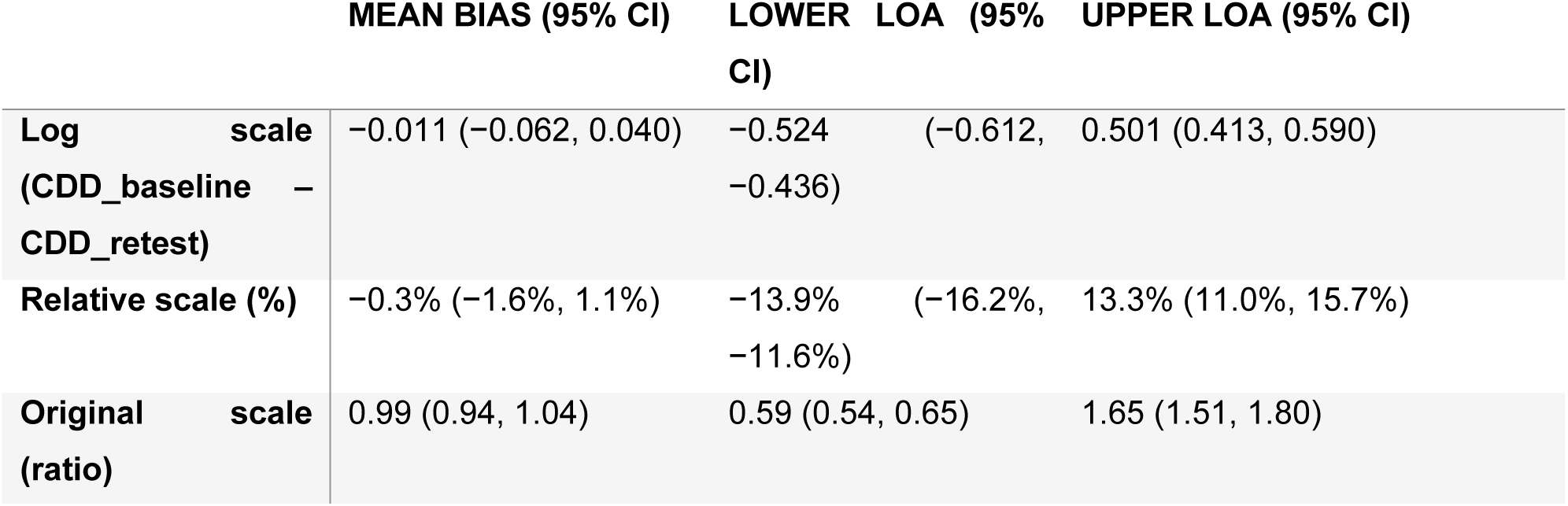
Bland-Altman Tests.

When expressed on the relative scale, the mean bias corresponded to −0.3% (95% CI: −1.6% to 1.1%), with limits of agreement from −13.9% to +13.3%. This indicates that, for most individuals, repeated measurements differed by no more than approximately ±14%.

On back-transformation to the original scale, the limits of agreement corresponded to CDD_baseline being approximately 0.59 to 1.65 times CDD_retest. In practical terms, this means that for most participants, the baseline CDD estimate was between 41% and 65% lower or higher than the retest estimate. Visual inspection of the Bland–Altman plot (Fig 5) showed that differences were symmetrically distributed around zero, with no evidence of proportional bias across the range of CDD values.

**Fig 5:**
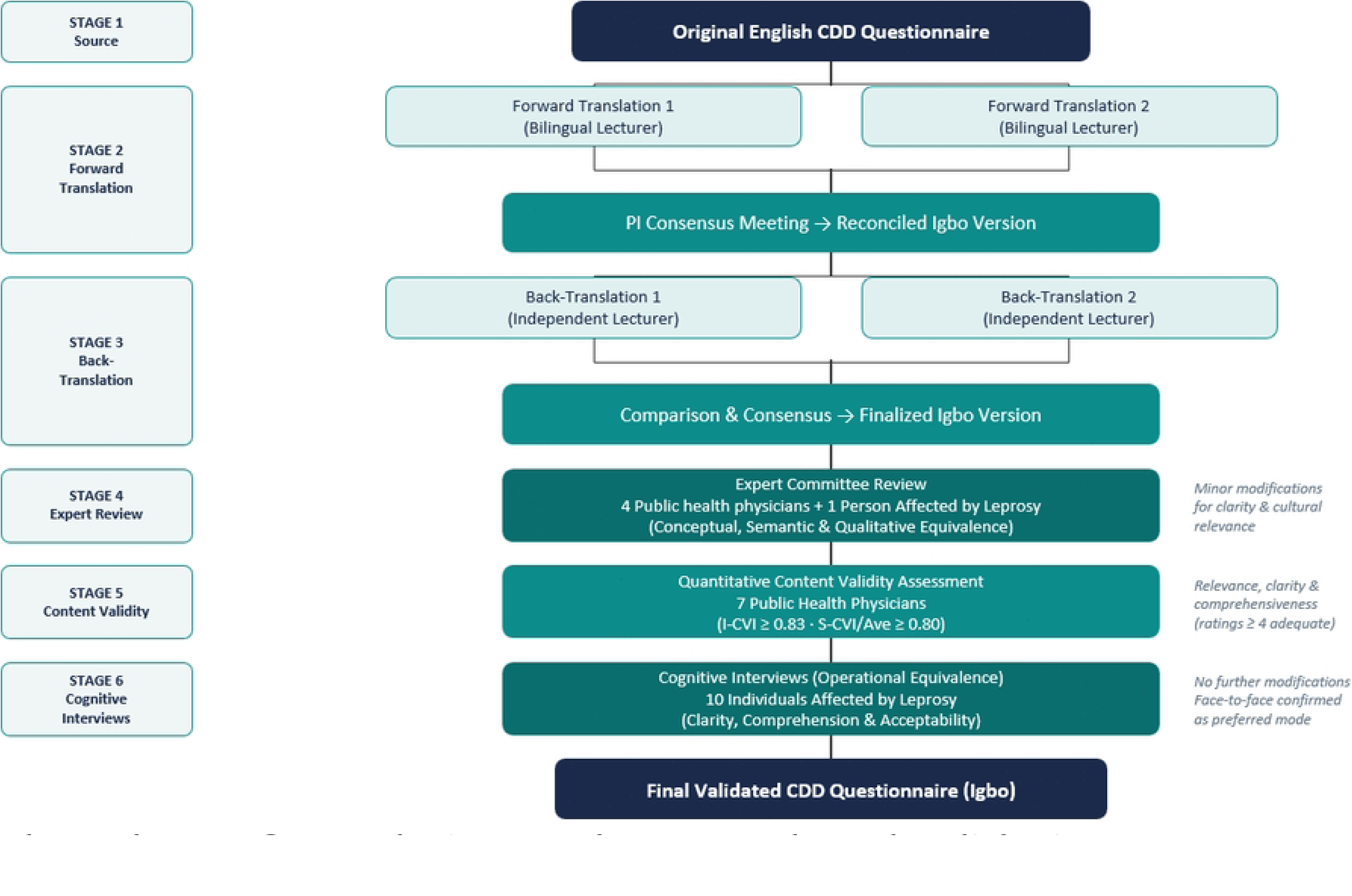
Bland-Altman plot of differences between CDD_baseline and CDD_retest.

## Discussion

Following systematic translation and cross-cultural adaptation, the instrument demonstrated good internal consistency, stability, content validity, and evidence of construct validity, supporting its suitability for leprosy research and program monitoring. These findings are consistent with validation studies of the same instrument conducted in Ethiopia, Mozambique, Tanzania, and Indonesia(11,12).

The observed inter-item correlations and internal consistency estimates indicate that the questionnaire items coherently measure the underlying construct of case detection delay. The pattern and magnitude of correlations among CDD components are theoretically consistent and provide evidence of construct validity, particularly in the absence of a gold standard or an established comparison tool for measuring diagnostic delay in leprosy.

Reliability assessment demonstrated excellent stability over time, with very high agreement between baseline and retest measurements (Spearman’s ρ = 0.985, p < 0.001) and an ICC for single measures of 0.996 (95% CI: 0.994–0.997). These results indicate that the instrument yields highly reproducible estimates of case detection delay and that a single measurement is sufficient for reliable assessment in this context.

Although the limits of agreement were relatively wide (–41% to +65%), this variability likely reflects the inherent skewness of delay data and the involvement of different raters at baseline and retest. Importantly, the absence of systematic bias, as indicated by a mean difference close to zero, suggests that the instrument produces consistent estimates on average.

This study represents the first cross-cultural adaptation and validation of the leprosy CDD questionnaire in Nigeria. The findings demonstrate that the questionnaire is a valid and reliable tool for estimating leprosy case detection delay in the Nigerian context. This is particularly important for the national leprosy control program, given the persistently high proportion of new leprosy cases presenting with G2D, which reflects late diagnosis.

Although the program currently attempts to capture case detection delay using a single data element (“duration of first symptoms (in months)” recorded on the leprosy treatment card), this approach is unlikely to yield estimates comparable to those obtained from the CDD questionnaire. The multiple, structured questions and recall prompts embedded in the CDD instrument facilitate more accurate reconstruction of key time points along the care-seeking pathway. Furthermore, reliance on a single variable does not permit separate estimation of patient delay and health system delay, both of which are critical for identifying bottlenecks and designing targeted interventions.

For a more comprehensive understanding of leprosy case detection delay, the CDD questionnaire should be complemented with additional questions that capture sociodemographic characteristics and other potential determinants of delay. This combined approach would enable more robust analyses to inform program planning and strengthen early case detection efforts in Nigeria and beyond.

## Limitations of the study

A limitation of this study is that the design did not allow for separate estimation of test–retest reliability (same rater across time) and inter-rater reliability (different raters at the same time). However, the very high ICC observed (>0.9) suggests that the instrument is robust to both sources of variability. Even when different raters assessed participants at different time points, the instrument produced highly consistent measurements, supporting its reliability for use in similar programmatic and research settings.

Reliance on self-reported timelines may be subject to recall bias, particularly given the long delays observed, although recall aids were used to mitigate it. Furthermore, the findings may not be fully generalizable to all regions of Nigeria, given regional heterogeneity in health system capacity and sociocultural practices.

## Data Availability

The data that support the study's findings are available on figshare at the following link https://figshare.com/s/b6a2d6ade2046b5bf0e0

## Acknowledgements

We sincerely appreciate the Royal Society of Tropical Medicine and Hygiene (RSTMH) providing funds for this project through the Early Career Research Grant program. We also gratefully acknowledge the invaluable support provided by the National Tuberculosis, Leprosy, and Buruli Ulcer Control Program, the State Tuberculosis, Leprosy, and Buruli Ulcer Control Program Managers, and the Local Government Tuberculosis, Leprosy, and Buruli Ulcer Control Program Supervisors. We also extend our heartfelt gratitude to all the individuals affected by leprosy who participated in this research. Their willingness to share their experiences was fundamental to the success of this research.

## Authors’ contributions

CCE and NMO conceptualized the study, developed the research questions, and constructed the questionnaire. CCE, NMO, NE, CN, MN, AN, OE, MS, JC, and OU contributed to the cross-cultural validation process and data collection. CCE performed the data analysis, interpreted the results, and drafted the initial manuscript. All authors critically reviewed the initial draft, edited it, and approved the final version.

